# Impact of PCSK9 Inhibitors on Lipoprotein(a): A Meta-analysis and Meta-regression of Randomized Controlled Trials

**DOI:** 10.1101/2024.07.10.24310245

**Authors:** Frederick Berro Rivera, Sung Whoy Cha, John Vincent Magalong, Vincent Anthony Tang, Mary Grace Enriquez, Martha Gulati, Byambaa Enkhmaa, Neha Pagidipati, Nishant P. Shah

## Abstract

**Background:** Lipoprotein(a) [Lp(a)] has been independently associated with increased cardiovascular risk. We examined the effect of monoclonal antibody proprotein convertase subtilisin/kexin type 9 inhibitors (PCSK9i) on plasma Lp(a) levels across multiple clinical trials.

**Methods:** Studies were retrieved comparing the effect of PCSK9i vs. placebo on Lp(a) levels. The primary outcome was percent change in Lp(a) levels. Secondary outcomes included percent change in additional cholesterol markers. Factors associated with the treatment effect were determined by meta-regression analysis. Subgroup analyses were done to explore potential treatment effect differences based on comparator, PCSK9i type, treatment duration, and presence of familial hypercholesterolemia (FH).

**Results:** 47 studies with 67,057 patients were analyzed. PCSK9i reduced Lp(a) levels on average of -27% (95% CI: -29.8 to -24.1, p<0.001). Concurrent reduction in LDL-C, non-HDL-C, total cholesterol, triglycerides ApoB, ApoA-1, and increased HDL-C were also observed with PCSK9i use. Factors associated with the treatment effect included mean percent change in LDL-C (p=0.02, tau^2^=177.1, R^2^=0.00) and Apo-B (p<0.00, tau^2^=114.20, R^2^=1.42). Subgroup analyses revealed consistent treatment effect amongst comparators (vs. placebo: -27.69% (95% CI: - 30.85 to -24.54, p<0.00), vs. ezetimibe: -24.0% (95% CI: -29.95% to -18.01, p<0.00), type of PCSK9i, evolocumab: -29.35% (95% CI: -33.56 to -25.14, p<0.00) vs. alirocumab: -24.50% (95% CI: -27.96 to -21.04, p<0.00), and presence of FH: -25.63% (95% CI: -31.96% to -19.30, p<0.00 vs. no FH: -27.22% (95% CI: -30.34. to -24.09, p<0.00). Varying treatment effects were noted in the duration of treatment (12 weeks or shorter: -32.43% (95% CI: -36.63 to -28.23 vs. >12 weeks: -22.31% (95% CI: -25.13 to -19.49, p<0.00), p interaction <0.01.

**Conclusion:** PCSK9 inhibitors reduce Lp(a) levels by an average of 27%. Mean percent change in LDL-C and Apo-B were associated with treatment effect. PCSK9i also significantly reduced other atherogenic lipoproteins. Across multiple clinical trials, PCSK9i has a consistent effect of significantly lowering Lp(a) levels.

## INTRODUCTION

Lipoprotein(a) [Lp(a)] is associated with premature and aggressive atherosclerosis across multiple vascular beds (1, 2), along with an increased risk of cardiovascular events.(3, 4) Plasma Lp(a) level is genetically determined and elevated in 20% of the general population.(5) Levels >50 mg/dL (or >125 nmol/L) are associated with significantly increased cardiovascular risk, independent of LDL-cholesterol (LDL-C) level, and therefore Lp(a) is considered a risk enhancer in ACC/AHA and ESC/EAS guidelines.(5, 6) Furthermore, a 3.5 fold higher Lp(a) levels increases the risk of death from cardiovascular disease (CVD), myocardial infarction (MI), and urgent coronary revascularization by 16%.(7) (8)

Proprotein convertase subtilisin /kexin type 9 (PCSK9) inhibitors (PCSK9i) have been postulated to lower Lp(a) level by reducing its production and enhancing clearance.(9) Patients who have higher baseline plasma Lp(a) level tend to experience greater Lp(a) lowering with PCSK9i therapy, which is associated with fewer adverse coronary events.(7) In a post-hoc analysis of the ODYSSEY Outcomes trial, reduction in Lp(a) by alirocumab contributed to a reduction in major adverse cardiovascular events (MACE), independent of LDL-C lowering.(10, 11). However, whether PCSK9i therapy lowers Lp(a) consistently across multiple studies is not well established.

Our meta-analysis aims to quantify the extent of Lp(a) reduction that can be achieved with PCSK9i therapy across multiple clinical trials, focusing on the monoclonal antibody variants like alirocumab and evolocumab. In addition to determining the percent reduction in Lp(a) by PCSK9i, we also aim to identify factors that influence the degree of Lp(a) lowering, as well as better understand the potential treatment effect differences based on PCSK9i type, comparator, treatment duration, and presence of familial hypercholesterolemia (FH). Concurrent reduction in other atherogenic lipoproteins with PCSK9i use is also assessed.

## METHODS

This study was reported under the Preferred Reporting Items for a Review and Meta-Analysis (PRISMA)(12), and the checklist(13) was followed **(see Supplementary Figure 1 and Supplementary Table 1)**. Certainty of evidence was rated using the Grades of Recommendation, Assessment, Development, and Evaluation (GRADE) framework.(14) This study was registered in the International Prospective Register of Systematic Reviews (PROSPERO)(15), with the identification number CRD42022378644.

### Data Sources and Searches

The literature search was performed using PubMed/MEDLINE, Ovid/Embase, Web of Science, SCOPUS, and Cochrane databases from database inception until May 2024. Search terms included "PCSK9 inhibitor", "PCSK9 antibody", "Evolocumab", "Alirocumab", "Bococizumab", "AMG145", "Repatha", "REGN727", "SAR236553", "RN 316", "PF-04950615", “LY3015014”, “RG7652”, “Lipoprotein a”, “Lp(a)”, “randomized controlled trial”, “randomization”, “clinical trials”, “intervention studies” and synonyms. PCSK9i therapies that are not monoclonal antibodies, such as inclisiran, were not included. Citations of selected articles and any relevant studies that evaluated Lp(a) and LDL-C lowering using PCSK9i were reviewed. After removing duplicates, records were reviewed at the title and abstract level, followed by the screening of full text based on our study criteria.

### Study Selection

Eligible phase II or phase III, double-blind randomized controlled trials (RCTs) comparing treatment with monoclonal antibody PCSK9i with placebo and/or ezetimibe in adult patients aged 18 years and above were included. Moreover, the studies must have reported baseline Lp(a) level and Lp(a) reduction in mean percent change, and treatment duration had to be 8 weeks or longer. Additionally, mean percent change in LDL-C and Apo-B from the baseline must have been reported. Studies were excluded if (1) they did not report a control arm, (2) they reported absolute change, or (3) Lp(a) was reported as median instead of mean. Other agents such as bococizumab and inclisiran were excluded because cardiovascular outcomes are not well established with these agents. We also excluded RCTs with participants younger than 18, and those reporting interim or post hoc analysis. Cross-over trials were also excluded due to the nature of the outcomes considered. Review articles, case reports, letters to the editor, commentaries, proceedings, laboratory studies, and other non-relevant studies were excluded as well.

### Data Extraction

Key participant and intervention characteristics and reported data on efficacy outcomes were extracted independently by two investigators (JM and VT) using standard data extraction templates. Any disagreements were resolved by discussion or, if required, by a third author (FBR). Data on the following variables were extracted: first author’s name, year of publication, journal, study phase, interventional and control treatments, randomization method, analysis tool, number of randomized patients, and demographic and clinical data (e.g., age, sex). In case of uncertainties regarding the study data, we contacted the authors of the specific study for additional information. Quality assessment was performed independently by two review authors (JM and VT) using the Revised Cochrane risk-of-bias tool for randomized trials.

### Outcome Measures

The primary endpoint of this meta-analysis and meta-regression was percent change from baseline in Lp(a) levels. Secondary endpoints included mean percent change from baseline in LDL-C, non-high-density lipoprotein cholesterol (HDL-C), total cholesterol, triglycerides, high-density lipoprotein cholesterol (HDL-C), apolipoprotein B (Apo-B), and apolipoprotein A1 (Apo-A1). Additionally, subgroup analyses were performed for applicable studies on the (1) type of PCSK9i, (2) comparator (placebo versus ezetimibe), (3) duration of treatment, and (4) population (FH versus no FH).

### Bias Assessment

All included studies reported a central randomization process, and outcomes were objectively determined. The included studies reported all primary and secondary outcomes as pre-specified in their protocols, so the risk of bias for selective reporting was judged as low. Two authors (JM and VT) independently assessed the risk of bias based on the Cochrane Risk of Bias Tool **(Supplementary Figures 2a and 2b)** for studies that fulfilled the inclusion criteria. Disagreements between the two reviewers were resolved by consensus. In case of persistent disagreement, arbitration by a third reviewer (FBR) was performed.

### Statistical Analysis

Stata Statistical Software version 18, College Station, TX: StataCorp LLC was used to conduct the included studies’ meta-analysis, heterogeneity tests, and sensitivity analyses. For all outcomes, the significance level was set at a p-value of <0.05. Statistical heterogeneity was identified through the forest plots and a standard Chi-squared test with a significant level of p<0.1. The extent of heterogeneity was based on the I^2^ statistic, wherein a value of more than 50% was interpreted as substantial heterogeneity. We pooled all estimates using a random effects model. Effect sizes were expressed using mean differences with 95% confidence intervals (CIs). Prespecified subgroup analyses were performed according to the (1) type of PCSK9i, (2) comparator (placebo versus ezetimibe), (3) duration of treatment, and (4) population (FH versus no FH). Regression analyses were performed to determine baseline factors such as LDL-C and Apo-B levels that could affect the point estimate. Changes in mean LDL-C was incorporated as possible covariate because of the reported higher discordance in LDL-C reduction for higher Lp(a) levels.(16) and Apo-B was incorporated in regression analysis as studies have suggested that it is not always linked on a single Lp(a) particle.(9)

## RESULTS

A literature search through July 2023, yielded 2,328 potentially relevant references on PCSK9i therapy, focusing on monoclonal antibody-based therapies (**Supplementary Figure 1**). Of these, 33 duplicates were removed. A total of 2,152 studies with unrelated interventions, outcomes, populations, non-original data (e.g., meta-analysis or review), descriptive or observational study design and study protocols were excluded. A total of 114 studies were left, and 35 pooled analyses were removed for not meeting the eligibility criteria. The remaining 79 related studies were retrieved as full-text publications for detailed evaluation. Overall, 47 studies were included in the final meta-analysis. From the 47 studies, data from 67,057 eligible individuals were included for analysis. The study characteristics are shown in **Table 1**.

**Table 1.**
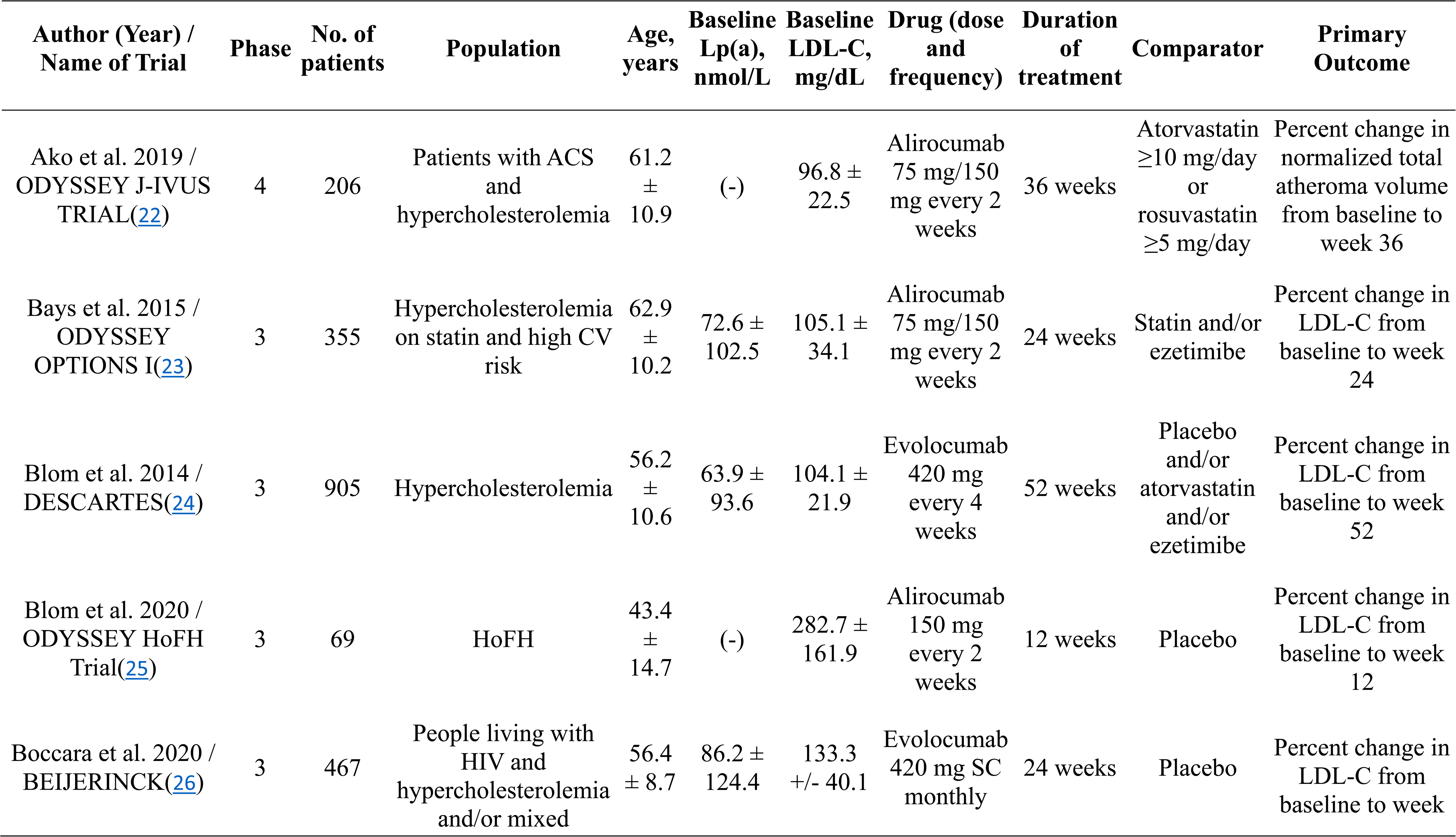
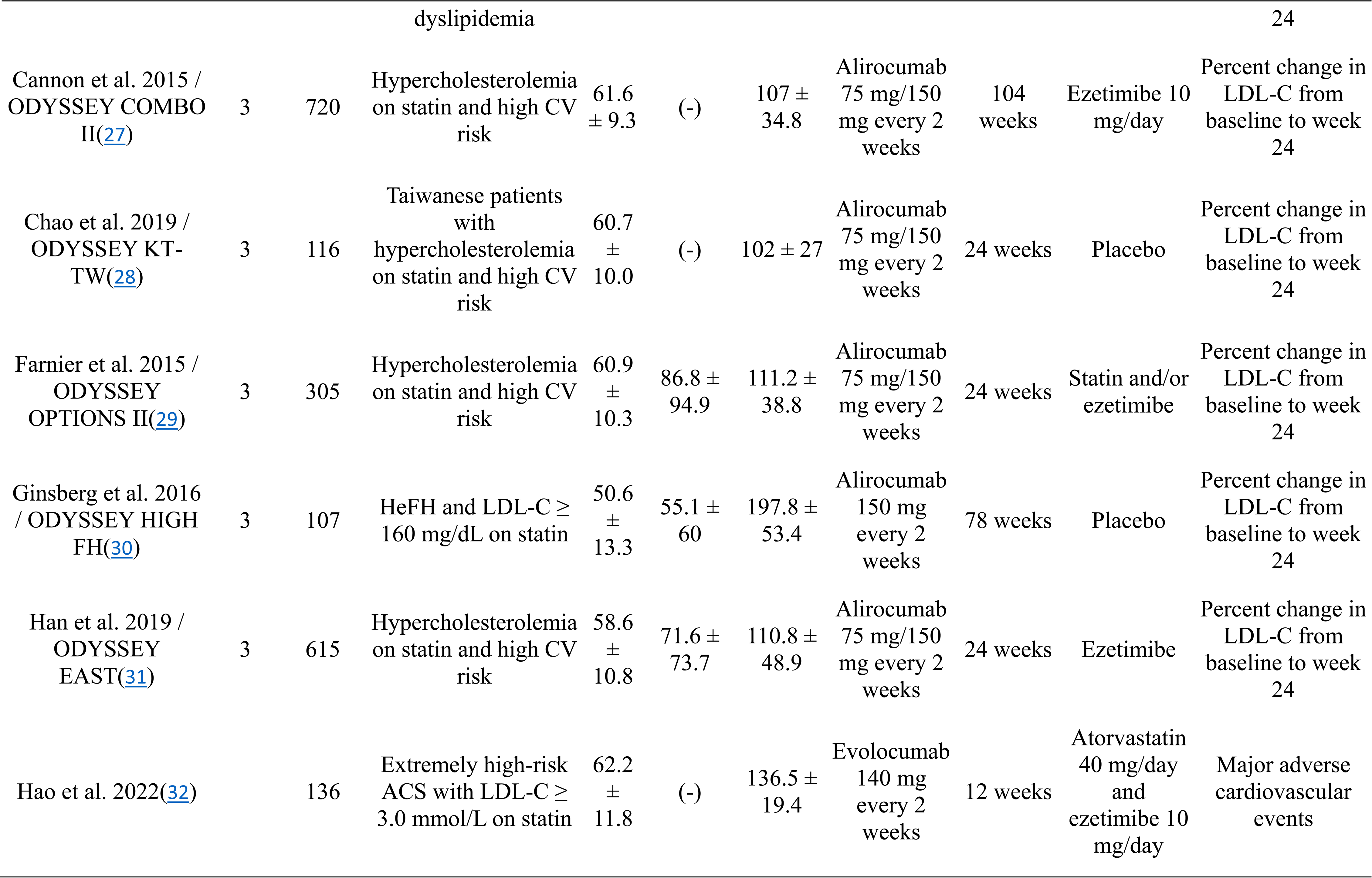
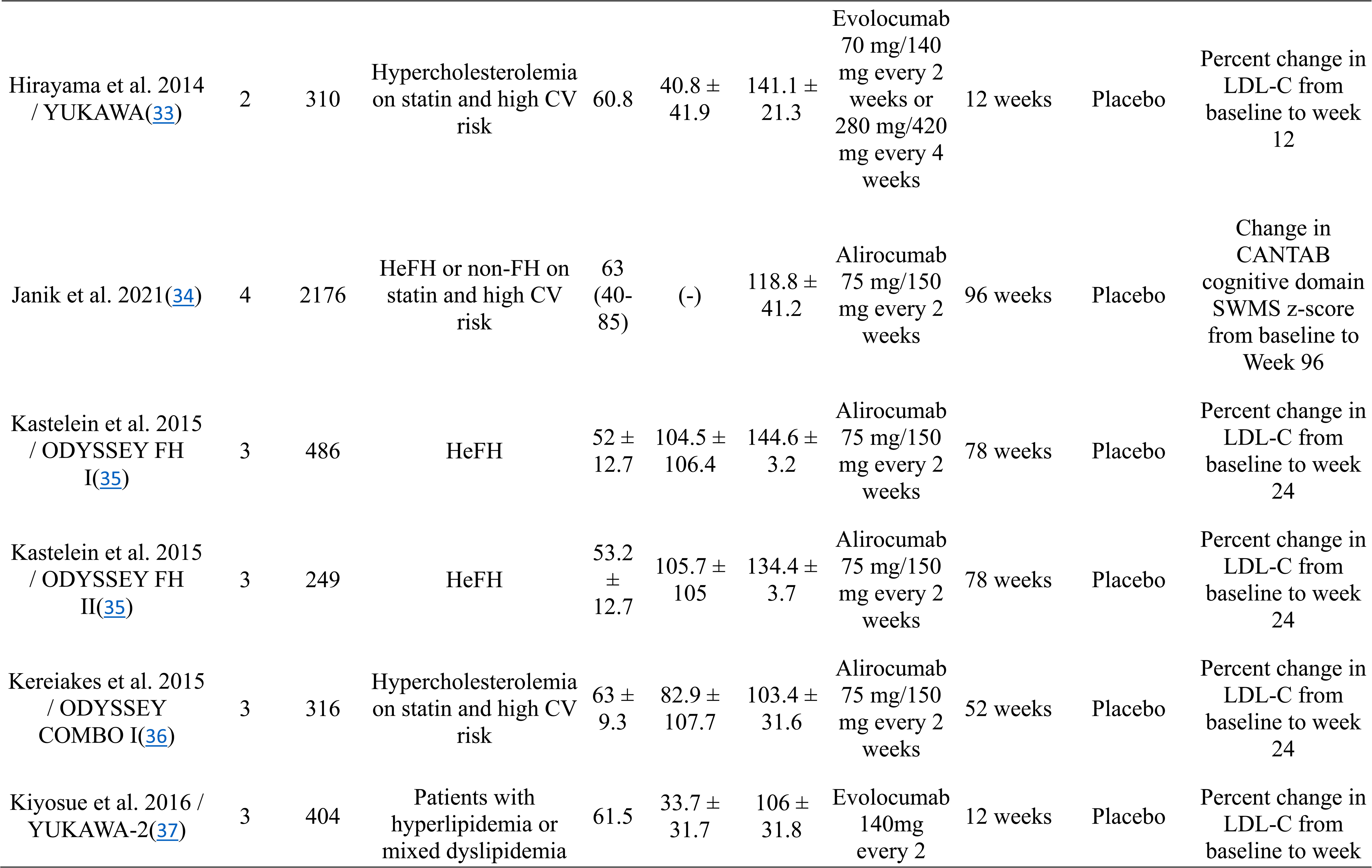
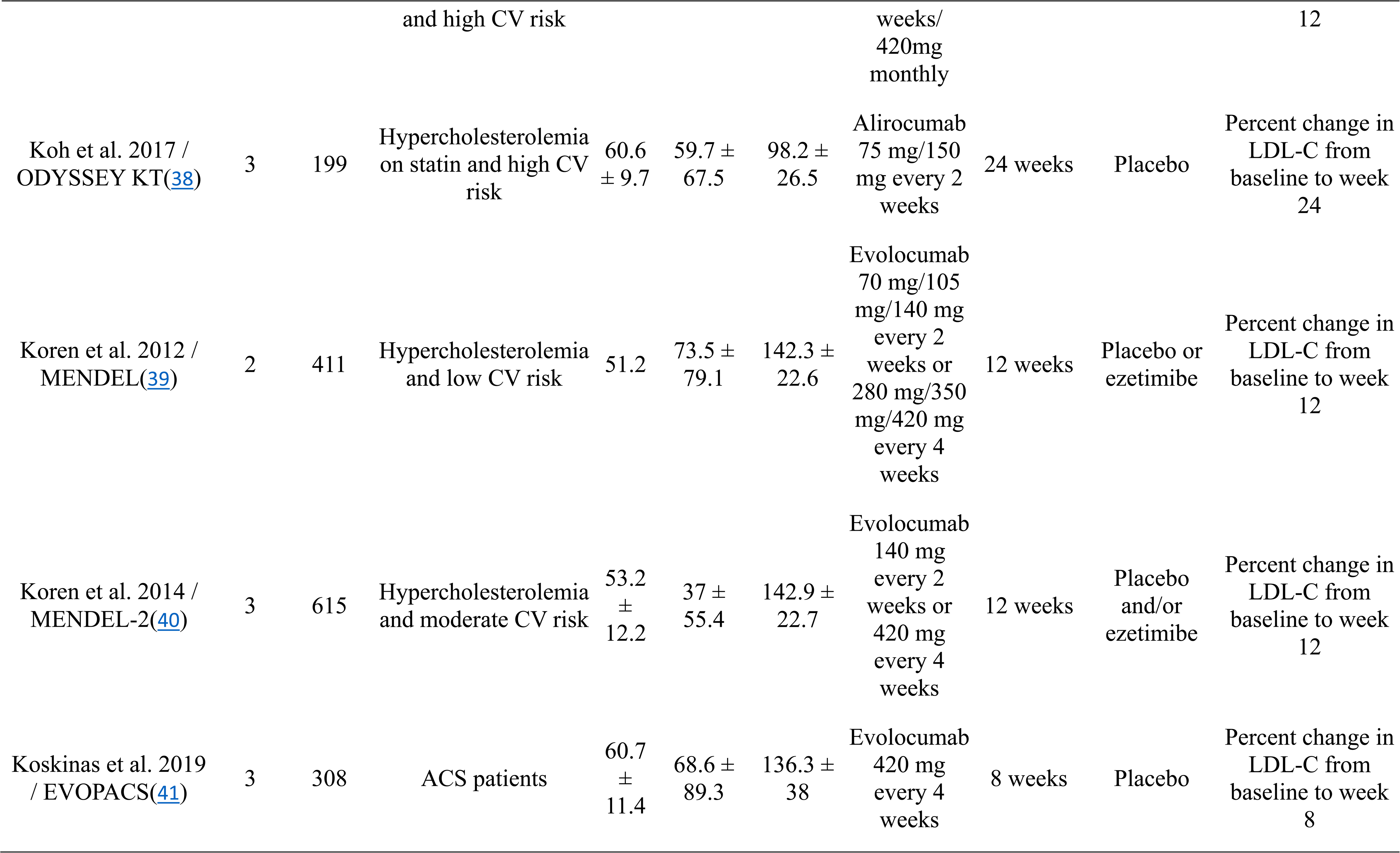
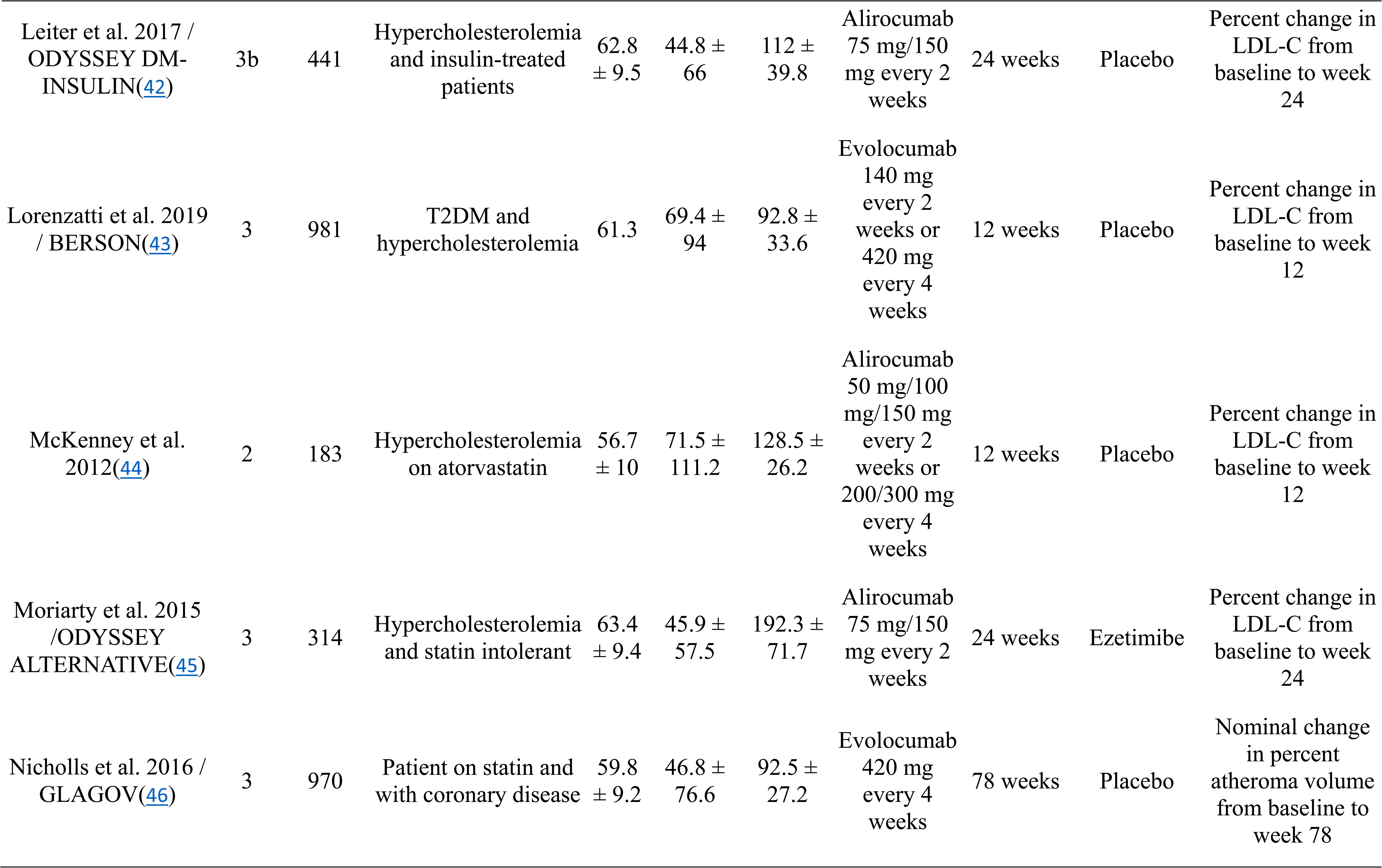
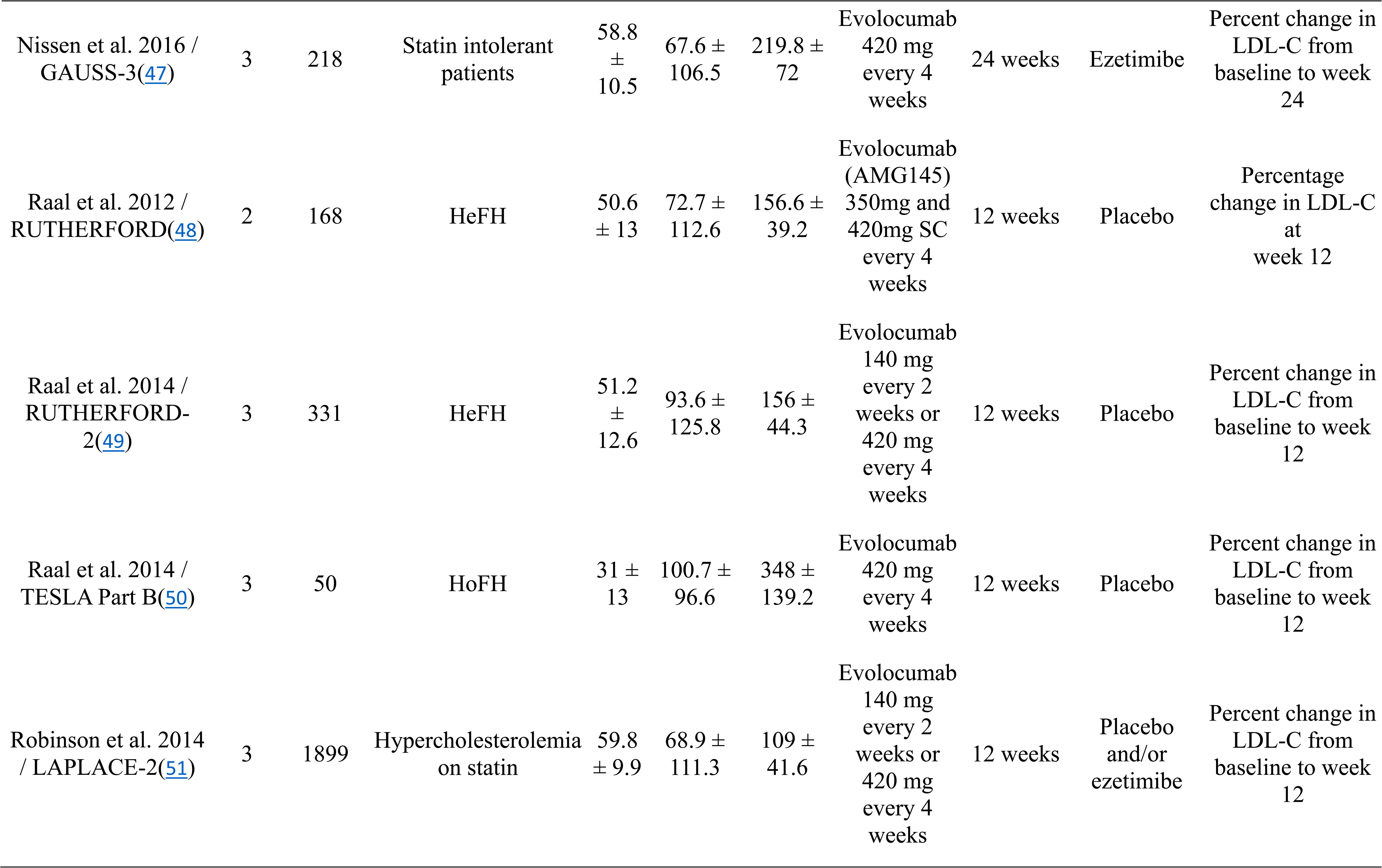
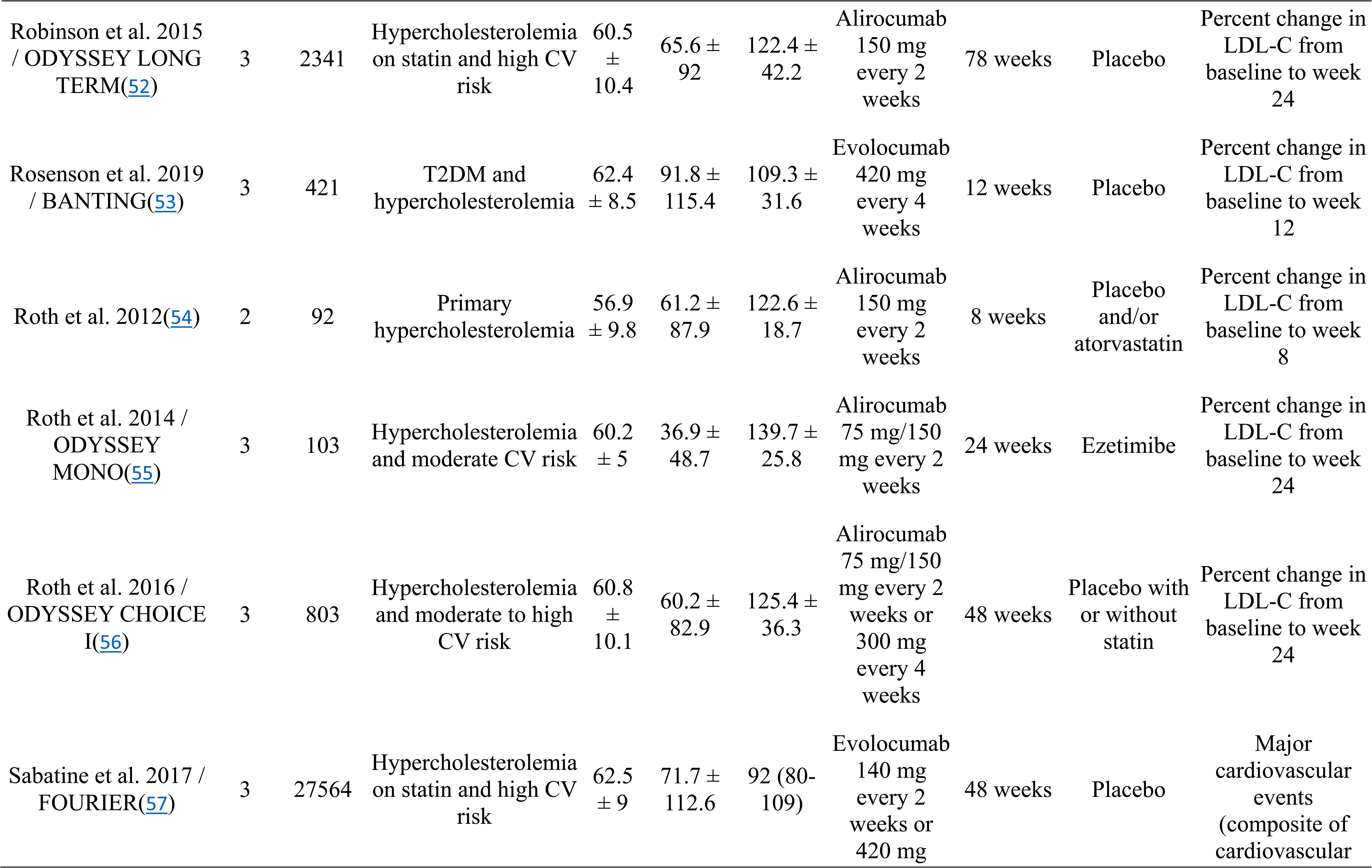
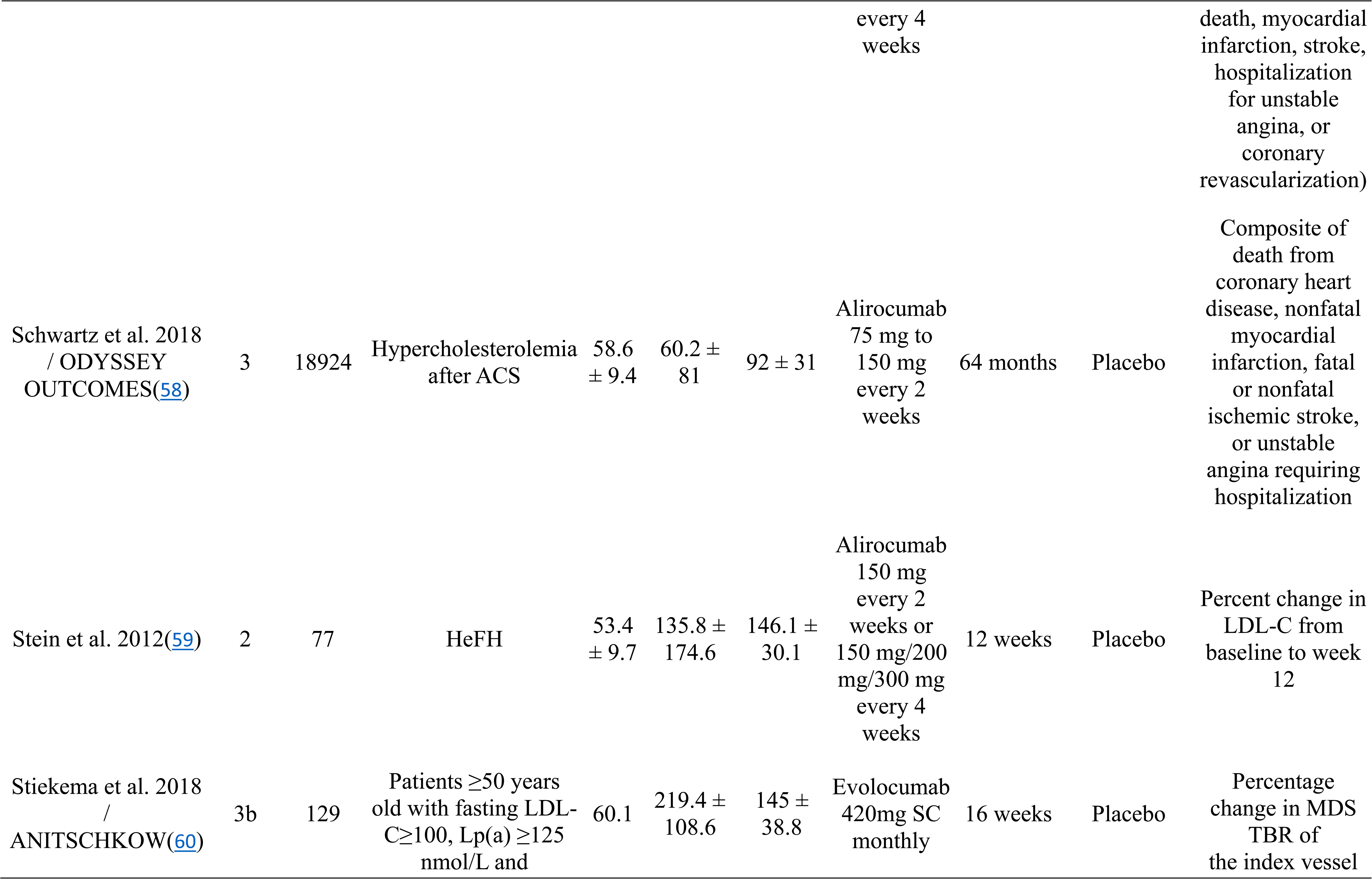
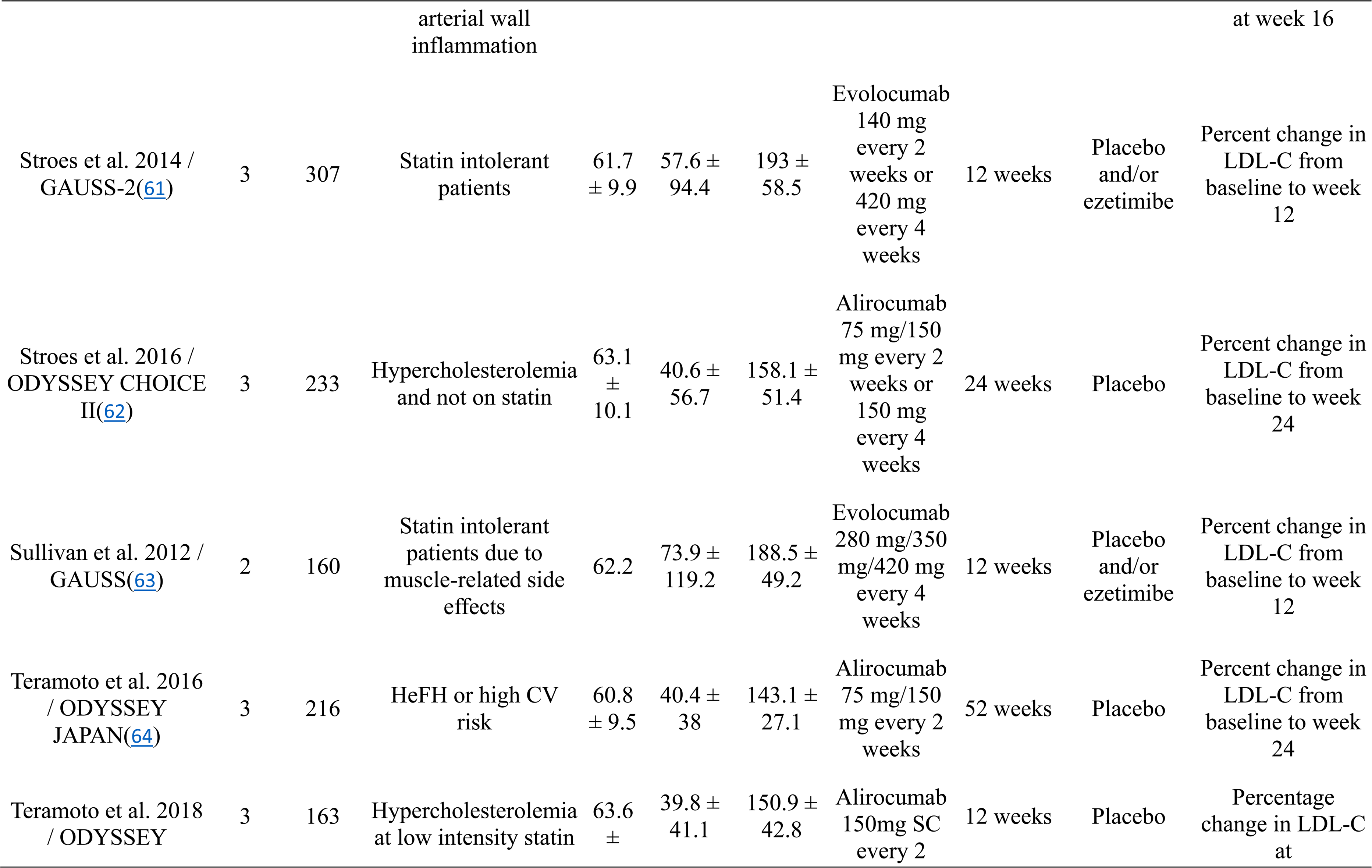
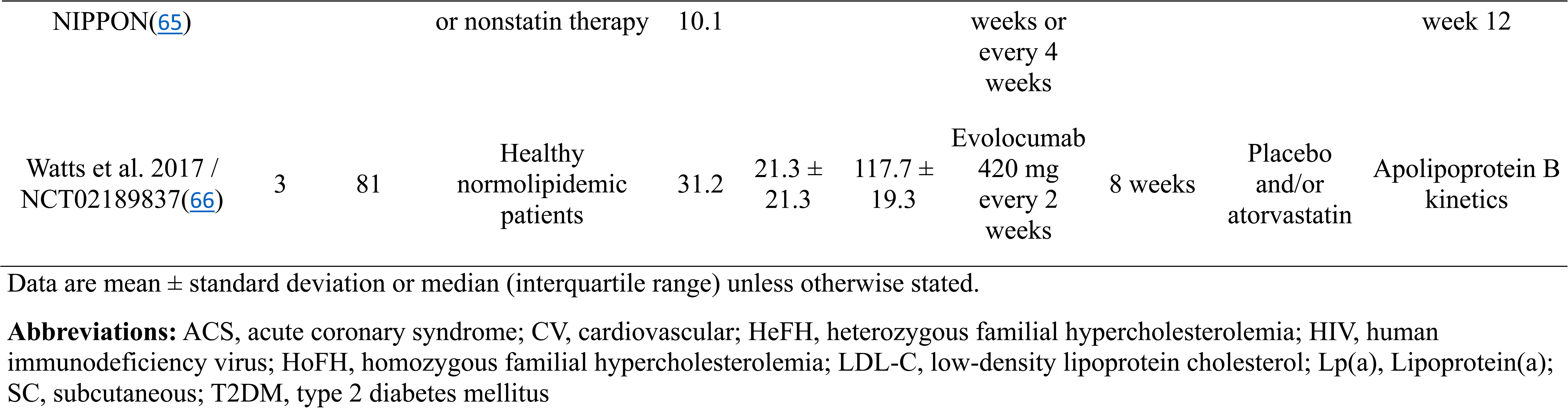
Characteristics of included studies.

### Lp(a) reduction

PCSK9i significantly reduced Lp(a) levels by -27% (95% CI: -29.8 to -24.1, p<0.00) on average compared with its comparator (placebo or ezetimibe). Significant heterogeneity was noted between the study results (I^2^ 96.16; p<0.00) **(Figure 1)**, which could be explained by the differences in patient risk profile and population characteristics (i.e., comorbidity like FH, race), type of PCSK9i, and treatment duration among others.

**Figure 1:**
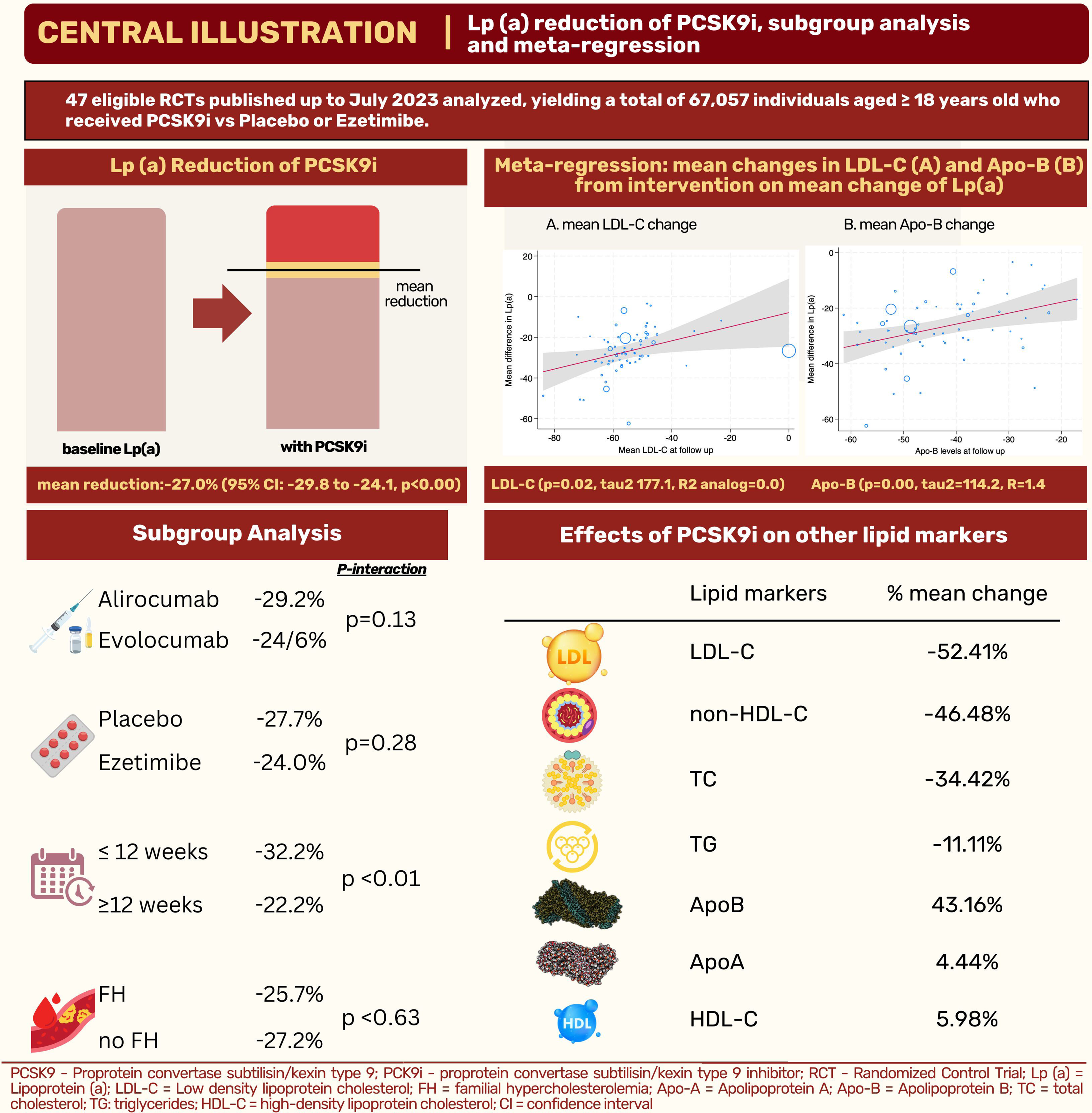
Central Illustration

### Subgroup analyses

PCSK9i significantly reduced Lp(a) by -28% when compared to placebo mean difference (MD) - 27.69% (95% CI: -30.85 to -24.54), p<0.00], and -24% when compared to ezetimibe [MD - 24.0% (95% CI: -29.95% to -18.01), p<0.00]. There was no significant difference in the treatment effect of PCSK9i compared with ezetimibe or placebo (p interaction 0.25) (**Figure 2**). The treatment effects of alirocumab and evolocumab on Lp(a) lowering were not significantly different (p interaction 0.06) (**Figure S3).** The achieved Lp(a) reduction by PCSK9i use was significantly different between those who had received treatment for equal to and less than 12 weeks [-32.43% (95% CI: -36.63 to -28.23)] vs. those who received treatment for more than 12 weeks [-22.31% (95% CI: -25.13 to -19.49, p<0.00)] (p interaction <0.01) (**Figure S4).** PCSK9i reduced Lp(a) for both FH and non-FH cohorts. (**Figure S5)**

**Figure 2:**
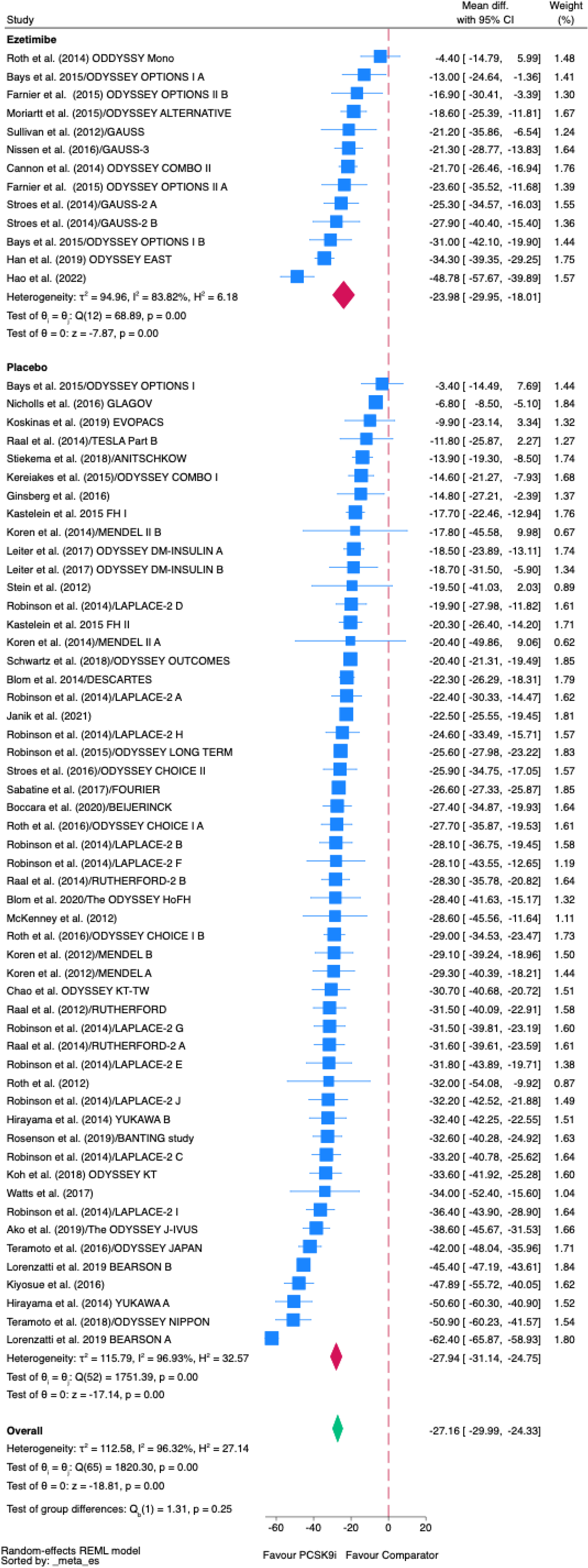
Forest plot on the effect on PCSK9i on lipoprotein a

### Meta-regression analysis

Meta-regression analysis revealed that the mean change of LDL-C (p=0.02, tau^2^=177.10, R^2^ analog=0.00) and Apo-B (p<0.001, tau^2^=114.20, R=1.42) were associated with the effect size difference **(Figures 3 and 4)**. Baseline LDL-C (p=0.14, Q=2.17, R^2^ analog= 0.00) and baseline mean age (p=0.43, Q=0.63, R^2^ analog=0.00) were not associated with the treatment effect.

**Figure 3:**
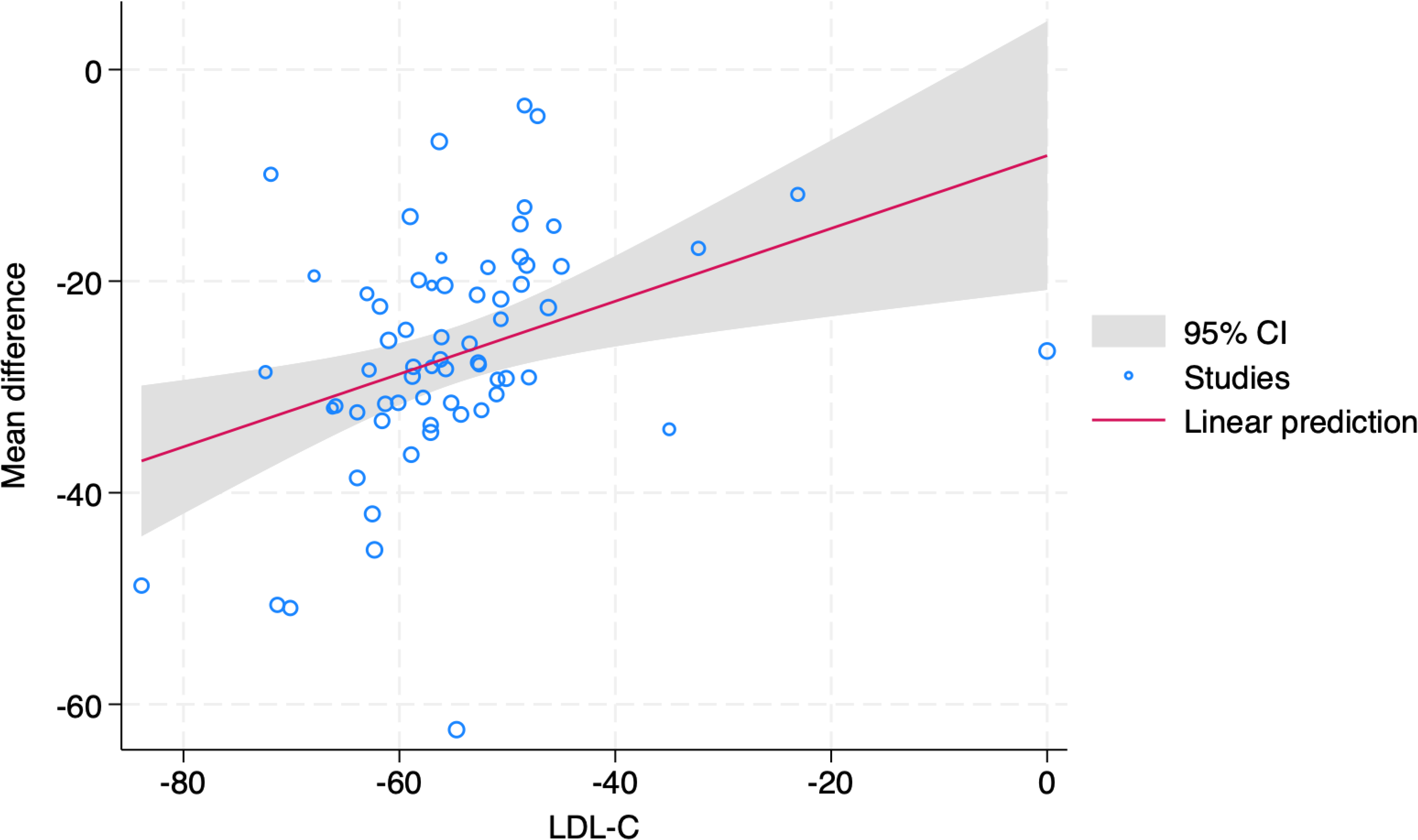
Forest plot on the effect sizes on different prespecified subgroups

**Figure 4:**
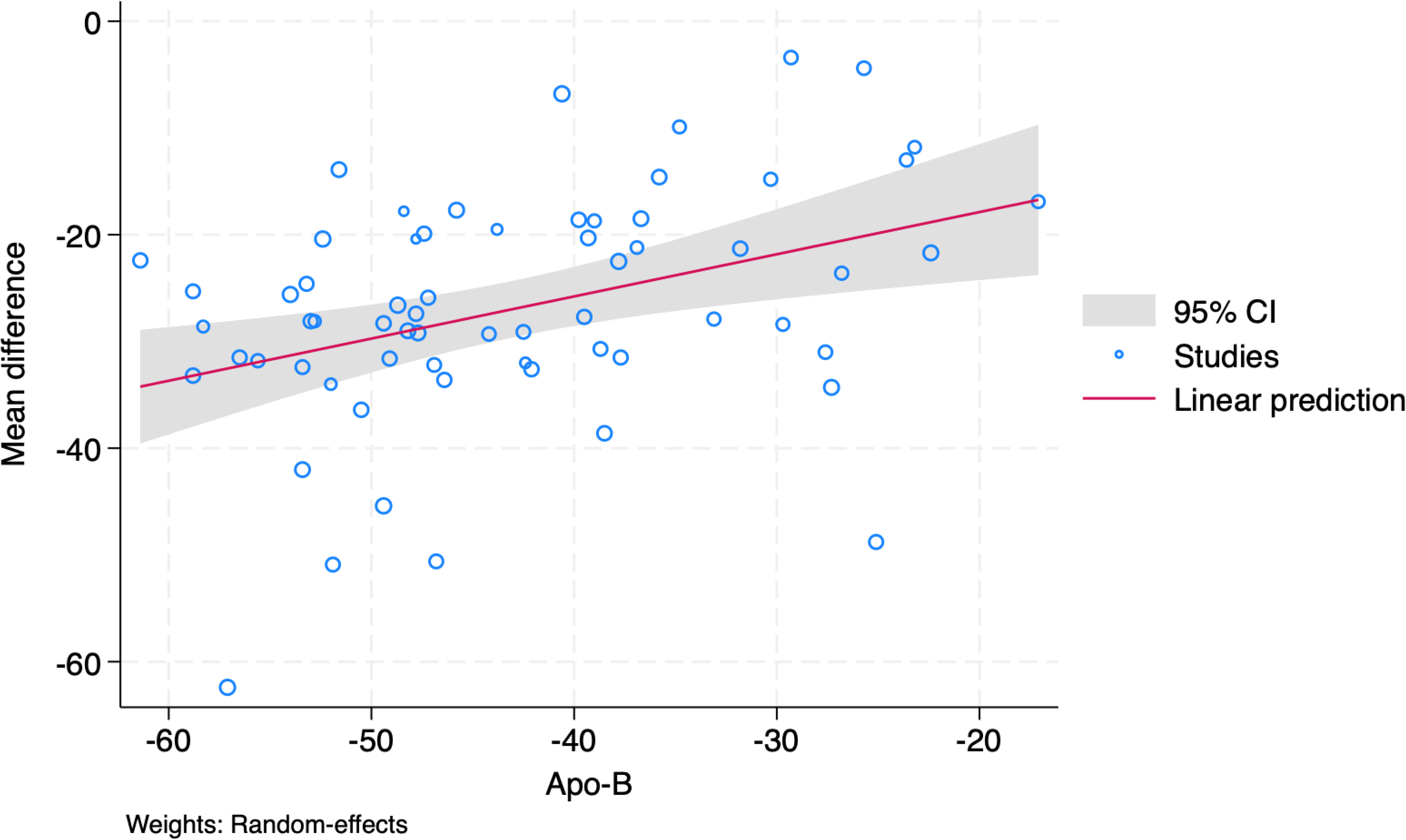
Scatter plot on the meta-regression analysis of mean changes in LDL-C (A) and Apo-B (B) from intervention on mean change of Lp(a)

### Effects of PCSK9 inhibitors on other lipid markers

PCSK9i reduced LDL-C by -52.41% [(95% CI -55.28 to -49.54%), p<0.01]. PCSK9i also reduced non-HDL-C by -46.48%, TC by -34.42%, triglycerides by -11.11%, and Apo-B by 43.16%. PCSK9i increased ApoA-1 by 4.44% and HDL-C by 5.98% (**Figures S6-12**). **Table 2** shows the summary results on the effects of PCSK9i on other lipid markers.

**Table 2:** Summary on the effects of PCSK9i on other lipids.

| **Lipids** | **Overall mean percent change** | **Mean percent change compared to placebo** | **Mean percent change compared to ezetimibe** |
| --- | --- | --- | --- |
| **LDL-C** | 52.41%<br>95% CI -55.28 to -49.54%, p<0.001 | -57.51%<br>95% CI -59.53 to -55.49, p<0.001 | -32.23%<br>95% CI -36.36 to -28.1, p<0.001 |
| ***Non-HDL-C*** | -46.48%<br>95% CI -48.73 to -44.23, p<0.001 | -50.87%<br>95% CI -52.57 to -49.16, p<0.001 | -28.6%<br>(95% CI -31.62 to -25.49, p<0.001) |
| ***Total cholesterol*** | -34.42%<br>95% CI -36.09 to -32.75, p<0.001 | -36.31%<br>(95% CI -37.68 to -34.94, p<0.001) | 21.47%<br>(95% CI -25.58 to -17.37, p<0.001) |
| ***Triglycerides*** | -11.11%<br>95% CI -12.86 to -9.35, p<0.001 | -13.27%<br>(95% CI -14.84 to -11.70, p<0.001) | -2.05%<br>(95% CI -4.42 to -0.31, p=0.09) |
| ***ApoB*** | -43.16%<br>95% CI -45.32 to -41.00, p<0.001 | -46.34%<br>(95% CI -48.08 to -44.60, p<0.001) | -30.62%<br>(95% CI -36.0 to -25.15, p<0.001) |
| ***Apo-A*** | 4.44%<br>95% CI 3.62 to 5.26, p<0.001 | 4.44%<br>(95% CI 3.62 to 5.26, p<0.001) | 4.64%<br>(95% CI 2.68 to 6.60, p<0.001) |
| ***HDL-C*** | 5.98%<br>95% CI 5.20 to 6.77, p<0.001 | 6.19%<br>(95% CI 5.33 to 7.06, p<0.001) | 4.95%<br>(95% CI 3.16 to 6.75, p<0.001) |

**Abbreviations:** PCSK9i= proprotein convertase subtilisin/kexin type 9 inhibitors; LDL-C= low density lipoprotein C; non-HDL-C=non-high density lipoprotein C; ApoB = apolipoprotein B; ApoA1 = apolipoprotein A1; HDL-C= high-density lipoprotein C

## DISCUSSION

In this meta-analysis of the impact of PCSK9i on Lp(a) level, we found that PCSK9i significantly reduced Lp(a) level regardless of comparator (placebo or ezetimibe), duration of treatment, type of PCSK9i, or presence of FH. Additionally, the mean percent change from baseline of LDL-C and Apo-B were associated with treatment indicating that the degree of Lp(a) lowering by PCSK9i depended on how much LDL-C and Apo-B were lowered. **(Central Illustration)**

Among the subgroups analyses, only the difference in duration of treatment had significant bearing on the treatment effect. Although our study demonstrated that PCSK9i leads to a modest Lp(a) lowering, the treatment effect appears to be higher in the first 12 weeks vs beyond 12 weeks of treatment. Some potential mechanisms underlying this observation have been proposed. Prolonged use of PCSK9i may induce its own resistance, which is thought to be from (1) increased endogenous PCSK9 triggered by PCSK9i use, and (2) delayed PCSK9 clearance due to accumulation of monoclonal antibody-PCSK9 complexes.(17) However, more data is needed to determine the possible reason of this treatment difference.

Many prospective studies have reported the role of PCSK9i in reducing Lp(a) levels(18), and prior pooled analyses have supported the efficacy of these agents.(19, 20) The results of our meta-analysis showed that the use of PCSK9i resulted in statistically significant reductions in plasma Lp(a) levels vs. comparators (placebo and ezetimibe), further increasing our understanding of the effect of PCSK9i on Lp(a) level based on data from multiple clinical trials. Furthermore, this analysis enriches available evidence by providing detailed information on subgroups of interest, based on duration of treatment, comparator, type of PCSK9i and presence of FH. The 27% reduction in Lp(a) level with PCSK9i compared to comparators (placebo and ezetimibe) in this analysis is consistent with the reports in previous meta-analyses citing on avergae26% reduction.(19, 20) Beyond confirming this PCSK9i’s effect on Lp(a), we also performed robust subgroup analyses with emphasis on duration of treatment and FH status. Using meta-regression analysis, we were able to determine factors associated with the treatment effect such as percent change of LDL-C and Apo-B from baseline. Changes in mean LDL-C was incorporated as possible covariate because of the reported higher discordance in LDL-C reduction for higher Lp(a) levels.(16) Apo-B was incorporated in regression analysis as studies have suggested that it is not always linked on a single Lp(a) particle hence can affect Lp(a) levels after PCSK9i.(9)

In previous studies by Farmakis and Yu, an Lp(a) reduction between 25-35% led to a clinically significant coronary heart disease (CHD) risk reduction(19, 20), and our data shows that monoclonal antibody based PCSK9i reduces Lp(a) level in the same range (-27%). Although studies have shown that a higher degree of absolute Lp(a) lowering (at least 50 mg/dL or 105 nmol/L) may be needed for a significant CHD risk reduction.(21), data from ODYSSEY Outcomes trial subanalysis showed the contrary.(10) Even a 1 mg/dL reduction in Lp(a) was associated with a hazard ratio of 0.99 (95% CI 0.99 to 0.00; p=0.008).(10) Trials on cardiovascular outcomes with Lp(a) reduction using directly targeting Lp(a) therapies such as the Lp(a)HORIZON (NCT04023552) and OCEAN(a) Outcomes trial (NCT05581303) are still ongoing, but they have shown reductions in Lp(a) level in the range of 80‒95%. Findings in these trials will help better understand the optimal absolute reduction in Lp(a) level needed for effective risk reduction.

### Limitations

There are several limitations that should be noted. This is a study-level meta-analysis, and we could not access individual patient data. Additional limitations include heterogeneity in PCSK9i studies. While we attempted to explain potential reasons for the heterogeneity using subgroup analyses, the lack of disaggregated data precluded further analyses for the matter. Sex and race are important subgroups that were sought from the studies reviewed; unfortunately, our findings show a gross lack of sex disaggregated data to permit subgroup analyses. Future trials should explore these specific disaggregated variables to obtain a deeper understanding of how the treatment effect may vary among these subgroups. Publication bias may also be present, the extent of which could not fully be quantified. Nonetheless, every effort possible was made to limit bias by utilizing a robust analytical approach to adjust for potential moderators through subgroup analyses and meta-regression.

## Conclusion

An elevated level of Lp(a) is an independent highly prevalent risk factor for atherosclerotic cardiovascular disease. In this analysis of 47 RCTs comparing PCSK9i monoclonal antibody therapy vs placebo or ezetimibe, PCSK9 inhibitors reduced Lp(a) levels by 27% on average. Mean change from the baseline in LDL-C and Apo-B positively correlates with Lp(a) reduction. Given the modest but significant reduction in Lp(a) level, further research is needed to evaluate the impact of monoclonal antibody based PCSK9i on cardiovascular outcomes in patients with elevated Lp(a).

## Data Availability

All data generated or analyzed during this study are included in this article. Further inquiries can be directed to the corresponding author.

## Acknowledgement

None

## Statement of Ethics

Ethics approval for this paper is not required because this study is based exclusively on published literature. Patient consent was not needed as this study was based on publicly available data.

## Conflict of Interest Statement

MG: Consultant Fees/Honoraria: Esperion, Novartis, and Boehringer Ingelheim Research/Research Grants: Congressionally Directed Medical Research Program-Department of Defense (WARRIOR study). NP: Consulting/honoraria: Bayer, Boehringer Ingelheim, CRISPR Therapeutics, Eli Lilly, Esperion, AstraZeneca, Merck, Novartis, and Novo Nordisk.; Grants to institution: Alnylam, Amgen, Boehringer Ingelheim, Eggland’s Best, Eli Lilly, Novartis, and Novo Nordisk, DSMB for trials sponsored by J+J and Novartis.

## Funding Sources

This paper was not funded.

## Author Contributions

Frederick Berro Rivera, MD, Sung Whoy Cha, MD, John Vincent Magalong, MD, Vincent Anthony Tang, MD, Mary Grace Enriquez, MD: **Data curation**; Frederick Berro Rivera, MD, John Vincent Magalong, MD: **Formal analysis**; Frederick Berro Rivera, MD, Sung Whoy Cha, MD, John Vincent Magalong, MD, Vincent Anthony Tang, MD, Mary Grace Enriquez, MD; **Investigation**; Frederick Berro Rivera, MD, Sung Whoy Cha, MD, John Vincent Magalong, MD, Vincent Anthony Tang, MD, Mary Grace Enriquez, MD: **Methodology**; Frederick Berro Rivera, MD, John Vincent Magalong, MD; **Validation**; Martha Gulati, MD MS; Byambaa Enkhmaa, MD, PhD; Neha Pagidipati, MD, MPH; Nishant P. Shah, MD: **Writing - original draft;** Frederick Berro Rivera, MD; Sung Whoy Cha, MD**: Writing -review & editing;** Martha Gulati, MD MS; Byambaa Enkhmaa, MD, PhD; Neha Pagidipati, MD, MPH; Nishant P. Shah, MD

## Abbreviations

ApoA1: apolipoprotein A1
ApoB: apolipoprotein B
CHD: coronary heart disease
CVD: cardiovascular disease
FH: familial hypercholesterolemia
HDL: high-density lipoprotein
LDL-C: low-density lipoprotein cholesterol
Lp(a): lipoprotein(a)
MACE: major adverse cardiovascular events
MI: myocardial infarction
non-HDL-C: non-high-density lipoprotein cholesterol
PAD: peripheral arterial disease
PCSK9: proprotein convertase subtilisin/kexin type 9
RCTs: randomized controlled trials

## Supplementary materials

### Supplementary figures

**Figure S1:** Preferred Reporting Items for Systematic Reviews and Meta-analysis (PRISMA) flow diagram

**Figure S2:** Risk of bias assessment

**Figure S3:** Subgroup analysis on type of PCSK9i

**Figure S4:** Subgroup analysis on duration of treatment

**Figure S5:** Subgroup analysis on type of population

**Figure S6:** Subgroup analysis on race

**Figure S7:** Forest plot on LDL reduction

**Figure S8:** Forest plot on non-HDL C reduction

**Figure S9:** Forest plot on total cholesterol reduction

**Figure S10:** Forest plot on triglycerides reduction

**Figure S11:** Forest plot on Apo-B reduction

**Figure S12**: Forest plot on HDL-C reduction

**Figures S13**: Funnel plots showing the risk of bias on the effect of PCSK9i on Lp(a) (A), Low density Lipoprotein Cholesterol (B), High density Lipoprotein Cholesterol (C) and non-High density Lipoprotein Cholesterol (D), Total Cholesterol (E), Triglycerides (F), Apo-A1 (G), and Apo-B (H).

### Supplementary table

**Table S1**: PRISMA Checklist

## Notes

### Competing Interest Statement

The authors have declared no competing interest.

### Clinical Trial

N/A

### Funding Statement

No specific financial support was obtained to prepare this article.

### Author Declarations

This is a study level meta-analysis. Approval from IRB was not necessary.

